# *RAB32*-linked Parkinson’s disease: Deep phenotyping, MDSGene literature review, and application of SynNeurGe criteria

**DOI:** 10.1101/2025.06.03.25328628

**Authors:** Teresa Kleinz, Francesco Cavallieri, Max Borsche, Giulia Toschi, Franco Valzania, Valentina Fioravanti, Enza Maria Valente, Pierfrancesco Mitrotti, Micol Avenali, Simone Zittel, Rommi Born, Michele Matarazzo, Alessio Di Fonzo, Edoardo Monfrini, Mandy Radefeldt, Letizia Santinelli, Norman Griebner, Cholpon Shambetova, Max Brand, Carolin Gabbert, Cornelis Blauwendraat, Joanne Trinh, Christian Beetz, Peter Bauer, Norbert Brüggemann, Global Parkinson’s Genetics Program (GP2), Christine Klein

## Abstract

**Background:** The *RAB32* p.Ser71Arg variant is a novel cause of monogenic Parkinson’s disease (PD), for which detailed phenotypic information is currently scarce.

**Objectives:** To clinically and biologically characterize individuals with PARK-*RAB32* to gain insights into genotype-phenotype relationships, disease severity, and underlying pathology.

**Methods:** We conducted a literature review following the MDSGene database protocol, alongside detailed phenotyping of 11 PARK-*RAB32* patients and one prodromal individual from the Rostock International PD (ROPAD) study. In addition to comprehensive scale-based assessments, including olfactory testing, we obtained neuroimaging data and various biomaterials, and performed α-synuclein seeding assays (SAA) in cerebrospinal fluid in a subset.

**Results:** 83 patients (74 from the literature) were included in the analysis. The median age at onset was 54 (IQR: 46-61) years. Typical Parkinsonism with a favorable dopaminergic response was observed in all patients.

In our cohort, the mean MDS-UPDRS III score was 38.5±21.8 points. Autonomic symptoms were present in all individuals, and 10/11 patients had hyposmia. Several non-motor symptoms were reported for the first time in PARK-*RAB32*. Misfolded α-synuclein was identified in 2/2 patients, but not in the prodromal individual. 123I-FP-CIT imaging was available for eight patients, revealing neurodegeneration in all of them.

**Conclusion:** While PARK-*RAB32* is clinically and likely pathologically similar to idiopathic PD, our study underscores the importance of carefully assessing non-motor symptoms in this newly described form of PD. According to SynNeurGe criteria, PARK-*RAB32* is classified as S^+^ (evidence of synucleinopathy), N^+^ (neurodegeneration supported by imaging data), and G_F_^+^ (presence of a genetic variant).

## 1 Introduction

The recent discovery of the pathogenic variant p.Ser71Arg in the *RAB32* gene as a cause of Parkinson’s disease (PD)(1,2) is the first newly described monogenic form of PD in the last decade. Given their rarity and the present lack of deep phenotyping data, little is known about the distinct clinical and genetic characteristics of patients with PARK-*RAB32*, disease-related biomarkers, and the course of the disease(1–7). The p.Ser71Arg founder variant in *RAB32* leads to overactivity of the Leucine-rich repeat kinase 2 (LRRK2), encoded by a gene in which pathogenic variants were identified as the most frequent cause of monogenic PD(8). As in PARK-*SNCA*, PARK-*LRRK2*, and PARK-*VPS35*, the mode of inheritance in PARK-*RAB32* is autosomal dominant, with evidence of reduced penetrance(1).

Here, we provide deep phenotyping data, including a comprehensive assessment of non-motor symptoms, of eleven PARK-*RAB32* PD patients and one non-manifesting carrier of the *RAB32* p.Ser71Arg variant, including MR imaging and DaTSCAN data for a subset, and compare these findings with 72 *RAB32*-linked PD patients published to date by applying the standardized MDSGene protocol(9).

2024 marks the first attempts to classify PD biologically(10,11), with the main aim to target the molecular basis of PD even before its clinical manifestation. The SynNeurGe criteria that are based on the presence of α-synuclein, genetic status, and neurodegeneration, also embrace monogenic forms of PD(10). Given that synuclein is often not detected in PARK-*LRRK2*, which has a comparable mechanism, and that α-synuclein was not found in the single available autopsy report of a patient with PARK-*RAB32*(1), we partially fill this gap here by including the first cerebrospinal fluid (CSF)-based α-synuclein amplification assay (SAA) data and extending the SynNeurGe criteria to PARK-*RAB32*.

## 2 Methods

### Setting up and deep phenotyping of the PARK-*RAB32* cohort

Patients were identified within the Rostock International PD (ROPAD) Study(12), with the cohort expanded by recruiting additional family members. Phenotyping was conducted at the Institute of Neurogenetics and Department of Neurology, University of Lübeck, Germany, or off-site when patients were unable to travel. The Ethics Committee of the University of Lübeck (Ratzeburger Allee 160, Building 70, 23562 Lübeck) approved the study (2023-105), and we obtained written consent from all examined participants.

The diagnosis of PD was established using the Movement Disorders Society criteria(13). All participants underwent a detailed history taking, including family history, and a videotaped movement disorder examination, all conducted by the same movement disorder specialist (both on-site and off-site). Two experienced movement disorder specialists rated the clinical-neurological assessment, blinded to the genetic status. For this purpose, videos of healthy individuals, individuals with idiopathic PD, individuals with *RAB32-*PD, and first-degree relatives who were or were not carrying the Ser71Arg variant were used. The Hoehn and Yahr scale (H&Y)(14) and the MDS-Unified Parkinson’s Disease Rating Scale (MDS-UPDRS I-IV)(15) were used to assess disease severity, while the Montreal Cognitive Assessment (MoCA)(16) was employed to identify (mild) cognitive impairment with a cut-off score of 26 points. The State-Trait Anxiety Inventory (STAI)-Y1 and -Y2 (Spielberger *et al*., 1970) score (cutoff-value for anxiety: ≥40 points), the Geriatric Depression Scale (GDS; cutoff-value for depression: ≥5 points)(17), the REM sleep behavior disorder screening questionnaire (RBDSQ; cutoff-value for RBD: ≥5 points)(18), the Epworth Sleepiness Scale (ESS; cutoff-value for excessive daytime sleepiness: ≥ 5 points)(19), and the Questionnaire for impulsive-compulsive disorders in Parkinson’s Disease–Rating Scale (QUIP-RS)(20) were administered to detect other non-motor symptoms such as anxiety, depression, RBD, excessive daytime sleepiness, or impulse control disorders. Autonomic symptoms were evaluated using the Scales for Outcomes in Parkinson’s Disease - Autonomic Dysfunction (SCOPA-AUT)(21). Hyposmia was assessed using the University of Pennsylvania Smell Identification Test (UPSIT)(22), with percentiles determined using the Brumm et al. 2023 update for individuals >50 years of age (23). Individuals with a score below the 10^th^ percentile were considered hyposmic.

Seven participants underwent structural brain MRI to exclude concurrent brain lesions. As part of routine clinical care, FP-CIT SPECTs were performed in eight patients. The FP-CIT SPECT images were contributed by two different centers and acquired 4–5 hours after intravenous administration of 123I-FP-CIT (185 MBq DaTSCAN), which was injected 60 minutes after thyroid blockade with sodium perchlorate. All scans were evaluated through visual inspection as well as a semi-quantitative analysis using volumes of interest (VOIs), with the occipital cortex (OC) serving as the reference region for FP-CIT binding. Specific binding ratios were calculated for the entire striatum (Str/OC), the head of the caudate nucleus (Caud/OC), the whole putamen (Put/OC), and both the anterior (aPut/OC) and posterior (pPut/OC) segments of the putamen. Analyses were conducted separately for the left and right hemispheres.

### Genetic testing, haplotyping, and alpha-synuclein seed amplification assay (SAA)

Patients were previously enrolled in the ROPAD study(8), and other monogenic causes were excluded for all patients by panel-based sequencing of 68 genes related to PD or parkinsonism. Patients with available genome sequencing data from the ROPAD cohort (n=3,354 patients) who had either an early age at onset (AAO) or a positive family history were specifically tested for the *RAB32* p.Ser71Arg variant(12), and findings were confirmed by Sanger sequencing.

Haplotype analyses were performed using genome data and confirmed with long-range PCR, sequencing, and phasing by “genome walking”(24). Genetic ancestry was determined from whole-genome sequencing data by characteristic genotype-based clustering as described in Westenberger et al., 2024 (8).

Furthermore, a range of biomaterials was obtained, including serum, plasma, and CSF in a subset. The SAA to detect α-synuclein seed amplification in CSF was performed as described(25) at Amprion Inc.

### MDSGene literature review

We performed a literature search on the English-language literature using the PubMed database (https://pubmed.ncbi.nlm.nih.gov/), according to the standardized extraction protocol of MDSGene(9), and compared phenotypic descriptions of PD patients with the data from our PARK-*RAB32* cohort. The exact search term can be found in Supplementary Table 1. Until March 24, 2025, seven papers in the literature reported patients with PARK-*RAB32*(1–7). Here, we refer to the most recent or comprehensive descriptions of duplicate reports. Seven patients from our cohort were also previously reported(3,6), but we add deep-phenotyping, imaging, and SAA data.

## 3 Results

### Literature review and clinical comparison to our PARK-*RAB32* cohort

In the following, data from a combined study group of 83 PARK-*RAB32* patients (72 from the literature and 11 from our cohort) are presented. The median AAO was 54 years (IQR: 46-61; range: 31-82 years; missing data: 2.4%), the median age at examination (AAE) 70 years (IQR: 60.5-78; range: 44-98 years; missing data: 54.2%), and the median disease duration 14 years (IQR; 10-21; range: 0-39 years; missing data: 21.7%). Forty-six (55.4%) were women. Of the patients with (self-reported) ancestry information available (n=65; 78.3%), 87.7% were of European ancestry, 9.2% were of Tunisian ancestry, 1.5% Asian, and 1.5% of mixed ethnicity. A positive family history was reported in 74% of the patients with information available (n=77; 92.8%).

All included patients had classic parkinsonism with a good response to dopaminergic medication, with only one patient having atypical signs according to the MDSGene criteria (postural abnormalities in the sense of camptocormia and antecollis within the first ten years of disease onset) (Figure 1A). Detailed motor evaluation (H&Y, MDS-UPDRS III), as well as non-motor symptoms, were not reported in the literature in up to 98.6% of the patients (Figure 1B).

**Figure 1.**
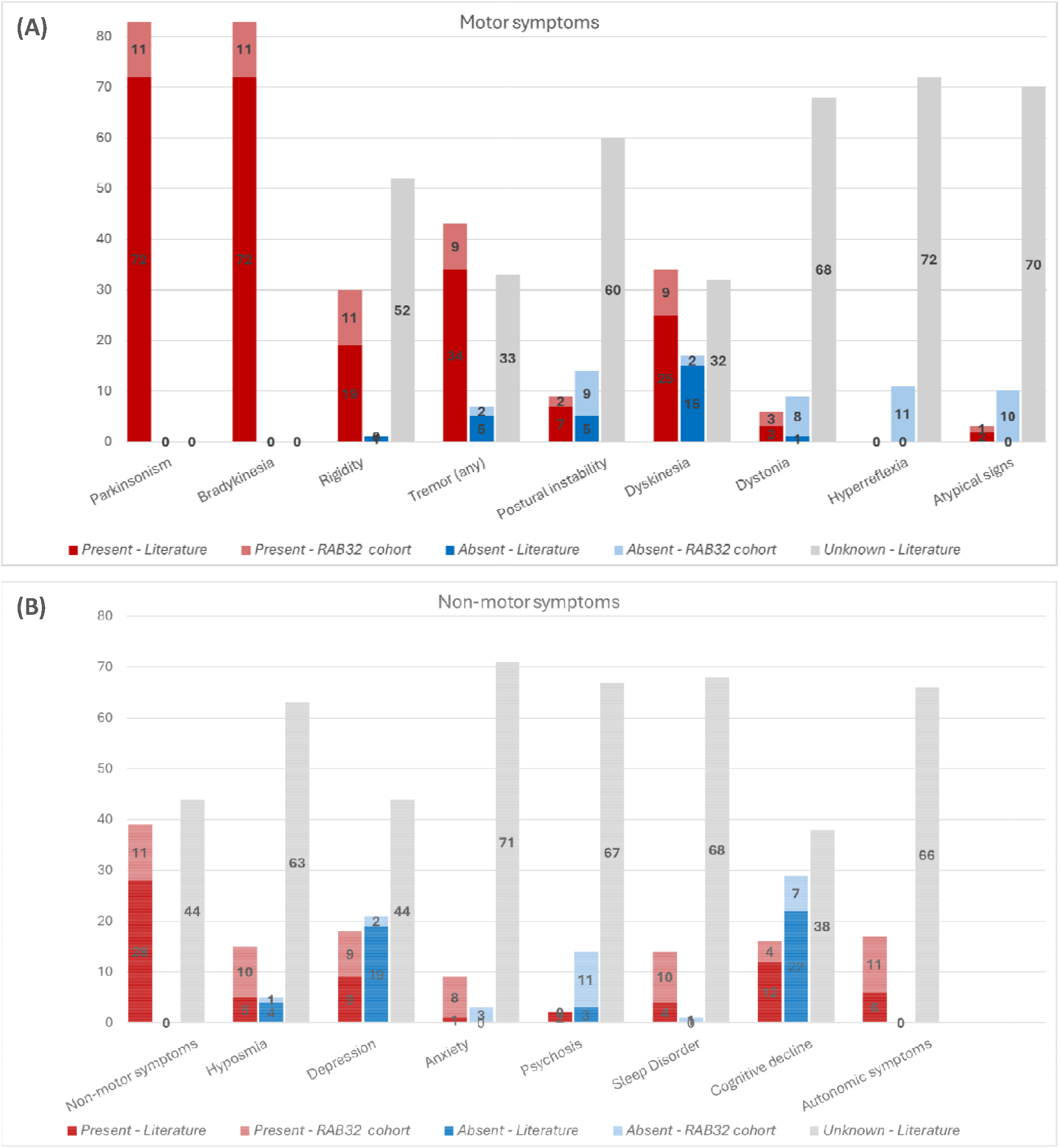
Presence of motor **(A)** and non-motor symptoms **(B)** in *RAB32*-PD, reported in the literature (dark red or blue parts of the stacked bar diagram) and observed in our PARK-*RAB32* cohort (light red or blue parts of the stacked bar diagram).

### Demographic, clinical, and genetic data in our PARK-*RAB32* cohort

From August 2024 to January 2025, we included twelve (10 women, 83%) individuals from ten distinct families with a heterozygous *RAB32* p.Ser71Arg pathogenic variant in our deeply phenotyped cohort. Eleven of them had a PD diagnosis, and one individual (who was the offspring of one of the patients) showed subtle motor signs compatible with PD but insufficient to establish a clinical PD diagnosis (Supplementary Video). Additionally, we examined two offspring of affected (female) PD patients who did not carry the pathogenic variant (Supplementary Table 2). 7/11 (63.6%) patients had a positive family history; Figure 2 shows two exemplary pedigrees displaying reduced penetrance. Nine patients were of Italian descent, one was Armenian, and one patient and her offspring originated from Germany. All were of (self-reported), and, in 5/5 individuals, genetically confirmed European ancestry (Supplementary Table 3), and all shared the same ancestral founder haplotype.

**Figure 2.**
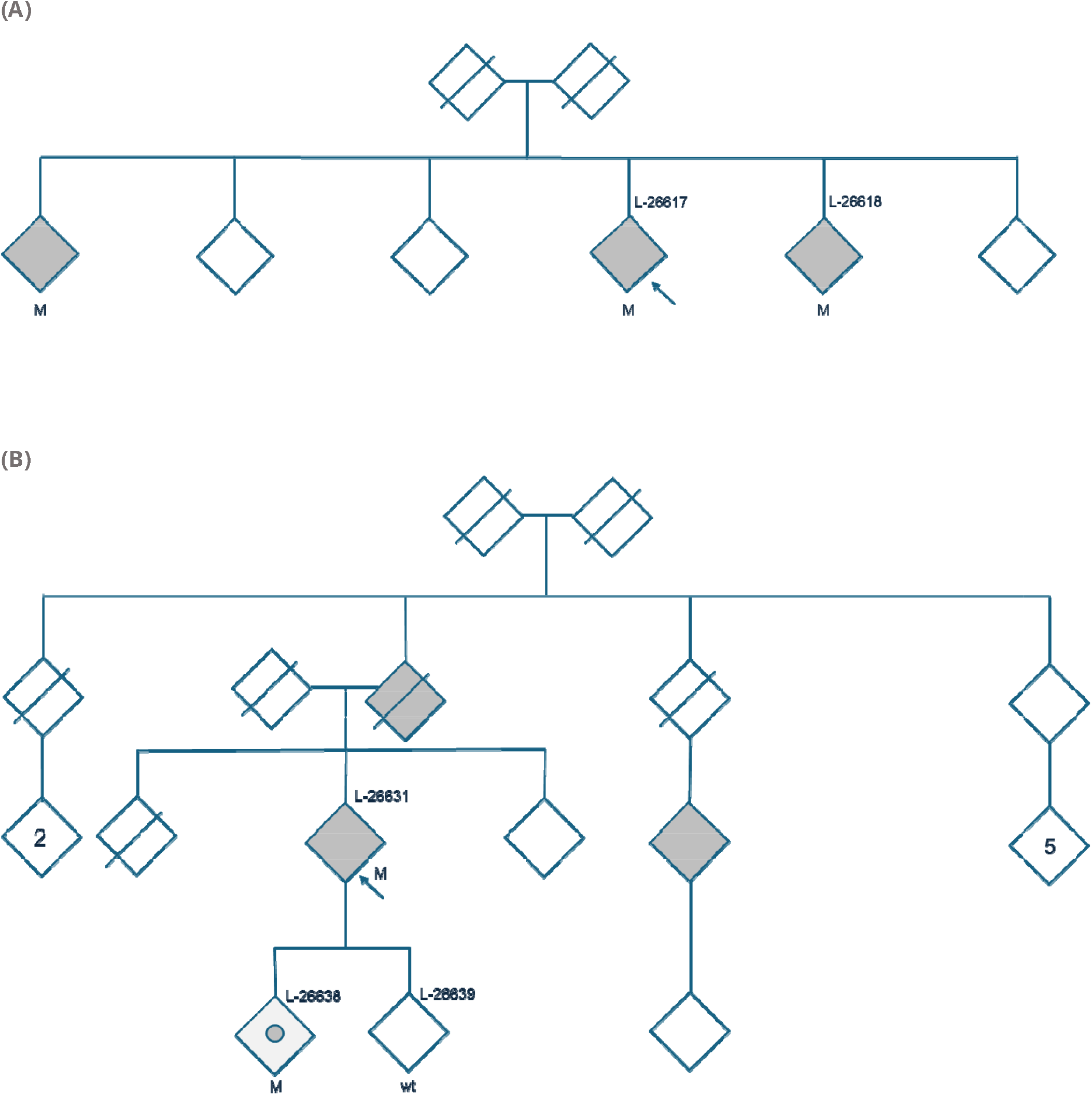
Pedigrees of *RAB32* families. The first family, ITA-I, from Italy **(A)**, comprises three siblings affected by *RAB32*-PD. Otherwise, the family history is blank. Both parents died in their late eighties. Two of the siblings were examined in this study (L-26617 and L-26618). The second family, GER-I, originates from Germany **(B)**. The Index patient (L-26631) and both offspring, L-26638 (*RAB32* p.Ser71Arg carrier with subtle signs) and L-26639 (wt), were examined. To further relatives from the maternal side are also affected by PD. PD, Parkinson’s Disease; M, confirmed mutant; wt, wildtype; AAO, age at onset; y, years

After a median disease duration of 11 years (IQR: 7-19.5), the median H&Y score in our cohort was 3 (IQR 2-3) points, the mean MDS-UPDRS III score was 38.5±21.8 points (ON-med), and the mean MoCA score was 23.2±6.1 points, with seven patients falling below the cut-off score of 26 points (Table 1, Supplementary Table 4, Supplementary Table 5). All patients self-reported motor fluctuations, while the mean MDS-UPDRS IV score showed 7.6±6 points (Supplementary Table 4). 9/11 (91.8%) patients had excessive daytime sleepiness upon ESS screening; 5/11 (45.5%) showed RBD, based on the applied questionnaire, and 8/11 (72.7%) rated above the cut-off for depression, according to the GDS. Hyposmia, scoring below the 10^th^ UPSIT percentile, was detected in 10/11 (90.9%) patients (more detailed individual-level data are available in Supplementary Table 4).

**Table 1.** Demographic and clinical aspects of the included individual *RAB32* carriers. All enrolled participants were heterozygous for the pathogenic variant p.Ser71Arg [G].

| Family ID | Local ID | Sex | AAO (~yrs) | DD (yrs) | H/Y | MDS-UPDRS III | MoCA | Seeding [S] | N within imaging* [N] |
| --- | --- | --- | --- | --- | --- | --- | --- | --- | --- |
| **Patients with PD** |  |  |  |  |  |  |  |  |  |
| ITA-I | L-26617 | m | 50's | 12 | 2 | 39 | 23 | yes | yes |
| ITA-I | L-26618 | m | 50's | 7 | 2 | 25 | 28 | N/A | yes |
| GER-I | L-26631 | f | 60's | 23 | 5 | 61 | 17 | N/A | N/A |
| ITA-II | L-26755 | f | 50's | 17 | 3 | 56 | 10 | N/A | N/A |
| ITA-III | L-26753 | f | 40's | 7 | 2 | 17 | 25 | yes | N/A |
| ARM-I | L-26809 | f | 50's | 8 | 2.5 | 12 | 25 | N/A | yes |
| ITA-IV | L-26914 | f | 60's | 11 | 3 | 36 | 27 | N/A | yes |
| ITA-V | L-26915 | f | 50's | 22 | 5 | 87 | 17 | N/A | N/A |
| ITA-VI | L-26934 | f | 60's | 3 | 2 | 31 | 30 | N/A | yes |
| ITA-VII | L-26913 | f | 40's | 7 | 3 | 34 | 28 | N/A | yes |
| ESP-I | L-27021 | f | 40's | 22 | 3 | 26 | 25 | N/A | N/A |
| ***RAB32* individual with subtle motor signs** |  |  |  |  |  |  |  |  |  |
| GER-I | L-26638 | f | 50's | N/A | 1 | 6 | 30 | no | N/A |

The individual with the *RAB32* pathogenic variant without an established PD diagnosis showed mild hypomimia and lateralized bradykinesia, including reduced arm swing and delayed shoulder shrug on the right side. In addition, depressive symptoms and constipation were reported to have been present for some years. Furthermore, RBD and excessive daytime sleepiness were identified (Supplementary Table 4). Examples of videotaped examinations are provided in the Supplementary Material.

### SAA, neuroimaging, and SynNeurGe criteria

CSF SAA revealed the presence of α-synuclein pathology in 2/2 *RAB32*-linked PD patients who donated CSF, while the individual with subtle signs of PD had a negative α-synuclein SAA result.

DaTScan results were available for 7/11 patients and confirmed neurodegeneration in all (Table 1, Supplementary Figure 1). The combined qualitative and quantitative analyses confirmed asymmetric tracer binding and a rostro-caudal gradient in six patients, consistent with the clinical picture of lateralized parkinsonism and the absence of significant asymmetry in one patient.

## 4 Discussion

For the first time, PARK-*RAB32* patients were classified according to the SynNeurGe classification criteria, including the evidence of synucleinopathy (S^+^), neurodegeneration based on imaging data (N^+^), presence of a genetic PD variant (G_F_^+^), in conjunction with the clinical picture of PD (C^+^)(10). These findings also serve as first evidence of the presence of misfolded α-synuclein in patients with PARK-*RAB32*, whereas the available single autopsy report described sparse neurofibrillary tangle pathology in the absence of α-synuclein pathology. Consistent with previous observations that hyposmia is a strong predictor of an underlying α-synuclein pathology (Simuni et al., 2024), both PD patients with hyposmia below the 10^th^ percentile for whom CSF was available showed positive SAA results, whereas SAA of the prodromal individual without hyposmia was negative. Some genetic PD forms do not consistently show Lewy body pathology in the brain and positive SAA in the CSF, which is mainly observed for patients with pathogenic variants in the autosomal recessive genes *PRKN* and *PINK1*, but also in PARK-*LRRK2* to varying degrees(26,27). The proportion of positive SAAs was reported to be 67.5% in PARK-*LRRK2*(27) and was especially present in patients or prodromal individuals with hyposmia, in keeping with the findings in our PARK-*RAB32* patients. In our sample, 10 out of 11 individuals exhibited hyposmia, which may suggest that PARK-*RAB32* is associated with a more prominent α-synuclein pathology compared to PARK-*LRRK2*, where hyposmia — and the associated α-synuclein pathology — is observed less frequently. Our findings also reveal possible pathophysiological implications, as RAB proteins are involved in the transport of α-synuclein, among other functions, whereby a defect can lead to maldistribution and ultimately to the promotion of aggregation(28).

The median AAO of PARK-*RAB32* was slightly lower than for PARK-*LRRK2* but higher than for autosomal recessive PD genes (Figure 3). However, we found a wide range of AAO from 31 to 82 years in the literature, calling for further investigations of the variable expressivity of the disease and modifying genetic and environmental factors. This observation should also be validated in larger cohorts and longitudinal studies of unaffected individuals with the *RAB32* pathogenic variant. Given the autosomal dominant inheritance pattern, the sex distribution in our cohort is more balanced compared to idiopathic PD, where a male predominance is typically observed.

**Figure 3.**
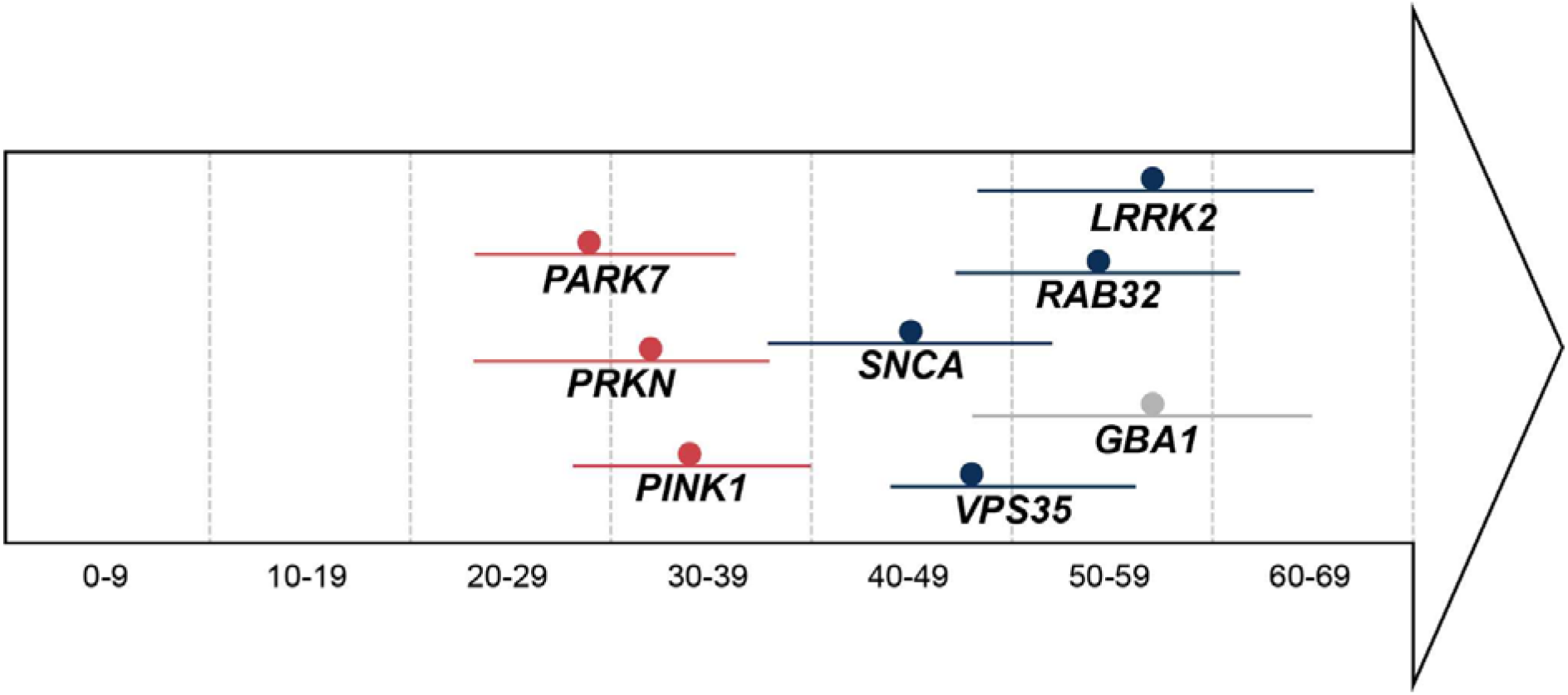
Comparison of the median age at onset. (and interquartile ranges) for patients with different genetic forms of Parkinson’s disease (PD), adapted from Vollstedt *et al*., 2023 (*Mov Disorders*) red: autosomal recessive inherited genes (*PRKN, PINK1, PARK7*) blue: autosomal dominantly inherited genes (*LRRK2, SNCA, VPS35, RAB32*) gray: *GBA1*

Remarkably, even in a newly discovered monogenic form of PD, the systematic literature search revealed a high proportion of unreported phenotypic features, especially non-motor symptoms. In our deeply phenotyped cohort, we identified a remarkably high prevalence of symptoms such as hyposmia, depression, anxiety, or autonomic dysfunction through targeted testing, highlighting the importance of detailed phenotyping and systematic reporting. In addition, no motor scales were reported (except for one person), which makes it difficult to draw conclusions about the severity and the individual course of the disease. A high proportion of missing data has also been found in previous MDSGene reviews for other monogenic forms of PD (29,30) and calls for systematic reporting of phenotype-genotype relationships. In addition, longitudinal data and further analysis of prodromal carriers are currently lacking in order to better understand disease progression.

Family history was negative in 26%, and pedigree analysis shows that some generations in larger families contain unaffected elderly individuals, indicating reduced disease penetrance. This is also known for individuals with PARK-*LRRK2*. Polygenic modulation has already been shown to be a mechanism influencing penetrance among individuals carrying the *LRRK2* p.G2019S variant and may also play a role in PARK-*RAB32*(31).

In conclusion, we i) identify the presence of alpha-synuclein seeding and apply the recent SynNeurGe criteria to PARK-*RAB32*; ii) demonstrate non-motor signs to be an important part of PD linked to *RAB32*; iii) provide the first MDSGene systematic literature review of PARK-*RAB32*.

## Data Availability

All data produced in the present study are available upon reasonable request.

https://amp-pd.org

https://www.mdsgene.org/

## 5 Declarations

### 5.1 Ethics approval and consent to participate

The Ethics Committee of the University of Lübeck (Ratzeburger Allee 160, Building 70, 23562 Lübeck) approved the study (2023-105), and we obtained written consent from all examined participants. The study was implemented in accordance with the ethical standards set out in the 1964 Declaration of Helsinki and its subsequent amendments.

### 5.2 Consent to publication

All patients signed informed consent to publication of the data collected.

### 5.3 Author Contributions

Teresa Kleinz Methodology, Formal Analysis, Investigation, Original Draft Preparation, Review and Editing

Francesco Cavallieri Methodology, Formal Analysis, Review and Editing

Giulia Toschi Methodology, Formal Analysis, Review and Editing

Franco Valzania Methodology, Formal Analysis, Review and Editing

Valentina Fioravanti Methodology, Formal Analysis, Review and Editing

Enza Maria Valente Methodology, Formal Analysis, Review and Editing

Pierfrancesco Mitrotti Methodology, Formal Analysis, Review and Editing

Micol Avenali Methodology, Formal Analysis, Review and Editing

Simone Zittel Methodology, Formal Analysis, Review and Editing

Rommi Born Methodology, Formal Analysis, Review and Editing

Michele Matarazzo Methodology, Formal Analysis, Review and Editing

Alessio Di Fonzo Methodology, Formal Analysis, Review and Editing

Edoardo Monfrini Methodology, Formal Analysis, Review and Editing

Letizia Santinelli Methodology, Formal Analysis, Review and Editing

Norman Griebner Methodology, Formal Analysis, Review and Editing

Cholpon Shambetova Methodology, Formal Analysis, Review and Editing

Max Brand Methodology, Formal Analysis, Review and Editing

Carolin Gabbert Methodology, Formal Analysis, Review and Editing

Cornelis Blauwendraat Methodology, Formal Analysis, Review and Editing

Joanne Trinh Methodology, Formal Analysis, Review and Editing

Mandy Radefeldt Methodology, Formal Analysis, Review and Editing

Christian Beetz Methodology, Formal Analysis, Review and Editing

Peter Bauer Methodology, Formal Analysis, Review and Editing

Norbert Brüggemann Methodology, Formal Analysis, Review and Editing

Christine Klein Methodology, Formal Analysis, Review and Editing

## 5.4 Acknowledgments

This project was supported by the Global Parkinson’s Genetics Program (GP2). GP2 is funded by the Aligning Science Across Parkinson’s (ASAP) initiative and implemented by The Michael J. Fox Foundation for Parkinson’s Research (https://gp2.org). For a complete list of GP2 members, see https://gp2.org.

This work was supported in part by the Intramural Research Program of the National Institutes of Health, including: the Center for Alzheimer’s and Related Dementias, within the Intramural Research Program of the National Institute on Aging and the National Institute of Neurological Disorders and Stroke. M.B. receives funding from the Else Kröner-Fresenius-Stiftung. J.T., N.B., and C.K. were supported by the DFG (FOR2488). N.B. was also supported by the EU Joint Programme - Neurodegenerative Disease Research (JPND). J.T. is supported by the DFG Heisenberg grant.

We would like to thank our study coordinator, Madita Grümmer, and the student assistant, Kiara Sparolin, for their support with the patient visits and with the data documentation.

## 5.7 Availability of data and materials

The data generated here is available via the Global Parkinson’s Genetics Program (GP2; https://gp2.org). Specifically, these are Tier 1/Tier 2 data from GP2 release R10. Tier 1 data can be accessed by completing a form on the Accelerating Medicines Partnership in Parkinson’s Disease (AMP®-PD) website (https://amp-pd.org/register-for-amp-pd). Tier 2 data access requires approval and a Data Use Agreement signed by your institution. All other data used can be found on https://www.mdsgene.org/.

