## Supplementary File for "*RAB32*-linked Parkinson’s disease: Deep phenotyping, MDSGene literature review, and application of SynNeurGe criteria"

### Supplementary Material

**Supplementary Table 1.** Search term for the literature search for *RAB32* on PubMed

| | |
|---|---|
| *RAB32* | (ataxia OR ataxic OR cerebellar OR channelopathy OR dystonia OR dystonic OR parkinson* OR paroxysmal movement OR tremor OR myoclon* OR chorea OR choreo* OR choreatic OR spastic paraplegia OR spastic paraparesis OR HSP OR Strümpell* OR hyperkinetic OR "movement disorder" OR dyskinesia OR dyskinetic) AND (*RAB32*) AND ("english"[Language]) |

**Supplementary Table 2.** Comparison of two offspring of patients with PARK-*RAB32*

| Participants (local ID, sex) | L-26638, female | L-27022, male |
|---|---|---|
| Index patient of the family | L-26631 | L-26921 |
| Genetic status | *RAB32* (Ser71Arg) positive (het) | *RAB32* (Ser71Arg) negative |
| Age at examination (~yrs) | 50's | 40's |
| UPSIT result | normal | normal |
| RBDSQ score | 5 points | 5 points |
| Motor examination findings | Mild hypomimia and lateralized bradykinesia, including reduced arm swing and delayed shoulder shrug on the right side* | Reduced arm swing and delayed shoulder shrug on the left side, bradykinesia, and interruptions in the foot-tapping test* |

*Supplementary Videos

~yrs, years (no exact to avoid potential identification); UPSIT, University of Pennsylvania Smell Identification Test; RBDSQ, REM sleep behavior disorder screening questionnaire; het, heterozygous

**Supplementary Table 3**. Genetic ancestry of RAB32-PD patients with WGS data available (n=5)

| **Local ID** | **ctry** | **Geo-region** | **AHG** | **Afri-can** | **Cauca-sian** | **Dravi-dian** | **East-afri-can** | **Near-East** | **North-euro-pean** | **Sahul** | **Sino-tibeta n** | **South-euro-pean** | ***** |
|---|---|---|---|---|---|---|---|---|---|---|---|---|---|
| L-26617 | ITA | Europe | 0 | 0 | 0,1997 | 0,1201 | 0,0557 | 0,1806 | 0,1126 | 0,0566 | 0,0611 | 0,2137 | 0 |
| L-26618 | ITA | Europe | 0 | 0,0694 | 0,2385 | 0,1011 | 0 | 0,1919 | 0,1159 | 0,0507 | 0,0608 | 0,1718 | 0 |
| L-26631 | GER | Europe | 0,0402 | 0 | 0,148 | 0,112 | 0 | 0,0825 | 0,2734 | 0,0714 | 0,0509 | 0,2216 | 0 |
| L-26753 | ITA | Europe | 0 | 0 | 0,1845 | 0,1241 | 0,0531 | 0,1713 | 0,1596 | 0,0473 | 0,0516 | 0,2085 | 0 |
| L-26915 | ITA | Europe | 0 | 0 | 0,1974 | 0,119 | 0,0598 | 0,1453 | 0,2035 | 0,0661 | 0 | 0,209 | 0 |

***Amerindian, Arctic, Austronesian, EA, Northindian, Paleosiberian, SEA, Saami, Samaritan, Samoedic, Siberian, Uralic**

ctry, country; ITA, Italy; Ger, Germany

**Supplementary Table 4.** Clinical signs and symptoms of the examined individuals

| **Individual local ID** | L-26617 | L-26618 | L-26631 | L-26755 | L-26753 | L-26809 | L-26914 | L-26915 | L-26934 | L-26913 | L-27021 | L-26638 |
|---|---|---|---|---|---|---|---|---|---|---|---|---|
| MOTOR | | | | | | | | | | | | |
| Parkinsonism | yes | yes | yes | yes | yes | yes | yes | yes | yes | yes | yes | no |
| Bradykinesia | yes | yes | yes | yes | yes | yes | yes | yes | yes | yes | yes | yes |
| Tremor | yes | yes | yes | yes | yes | yes | yes | yes | yes | yes | yes | no |
| Rigidity | yes | yes | yes | yes | yes | yes | yes | yes | yes | yes | yes | no |
| Postural instability | no | no | yes | no | no | no | no | yes | no | no | no | no |
| H/Y | 2 | 2 | 5 | 3 | 2 | 2.5 | 3 | 5 | 2 | 3 | 3 | N/A |
| MDS-UPDRS III | 39 | 25 | 61 | 56 | 17 | 12 | 36 | 87 | 31 | 34 | 26 | 4 |
| Dyskinesia | M.I. | M.I. | M.I. | M.I. | M.I. | M.I. | no | M.I. | no | M.I. | M.I. | no |
| Dystonia | no | no | no | no | no | no | yes | yes | no | no | yes | no |
| Hyperreflexia | no | no | no | no | no | no | no | no | no | no | no | no |
| Atypical | no | no | no | no | no | no | yes* | no | no | no | no | no |
| Diurnal fluctuations | yes | yes | yes | yes | yes | yes | yes | yes | yes | yes | yes | no |
| Levodopa response | good | good | good | good | good | good | good | good | good | good | good | N/A |
| Motor fluctuations | yes | yes | yes | yes | yes | yes | yes | yes | yes | yes | yes | no |
| MDS-UPDRS-IV | 9 | 7 | 12 | 16 | 5 | 0 | 0 | 17 | 0 | 9 | 9 | N/A |
| NON-MOTOR | | | | | | | | | | | | |
| Olfactory dysfunction | yes | yes | yes | yes | yes | yes | no | yes | yes | yes | yes | no |
| UPSIT score | 10 | 24 | 9 | 12 | 29 | 18 | 28 | 15 | 23 | 17 | 15 | 36 |
| UPSIT Percentile [%] | **1** | **7** | **1** | **2** | **5.5** | **5** | 25 | **4** | **5** | **1** | **4** | 62.5 |
| Depression | yes | no | yes | yes | yes | yes | yes | yes | no | yes | yes | no |
| GDS score | 4 | 1 | **9** | **9** | **8** | **8** | **6** | **6** | 0 | **8** | **7** | 3 |
| Anxiety | yes | no | no | yes | yes | yes | no | yes | yes | yes | no | yes |
| STAI-Y1 | 42 | 40 | 41 | 51 | 46 | 36 | 36 | 47 | 50 | 41 | 22 | 46 |
| STAI-Y2 | 46 | 44 | 53 | 49 | 46 | 48 | 41 | 37 | 47 | 39 | 22 | 50 |
| Psychotic | no | no | no | no | no | no | no | no | no | no | no | no |
| Cognitive decline | yes | no | yes | yes | yes | yes | no | yes | no | no | yes | no |
| MoCA | 23 | 28 | 17 | 10 | 25 | 25 | 27 | 17 | 30 | 28 | 25 | 30 |
| Sleep disorder | yes | yes | yes | yes | yes | yes | yes | yes | yes | yes | no | yes |
| RBDSQ | 3 | **11** | **10** | **12** | 4 | **10** | 0 | **7** | 1 | 3 | 3 | **5** |
| ESS | **14** | **6** | **17** | **7** | **15** | 3 | **6** | **7** | **10** | **10** | 2 | **10** |
| Autonomic | yes | yes | yes | yes | yes | yes | yes | yes | yes | yes | yes | yes |
| SCOPA-AUT | 23 | 24 | 21 | 33 | 8 | 36 | 6 | 21 | 10 | 7 | 26 | 5 |

**bold values**, above cutoff within scores; H/Y, modified Hoehn & Yahr scale; MDS-UPDRS, Movement Disorder Society-Unified Parkinson's Disease Rating Scale; UPSIT, University of Pennsylvania Smell Identification Test; GDS, Geriatric Depression Scale; State-Trait Anxiety Inventory, STAI; MoCA, Montreal Cognitive Assessment; RBDSQ, REM sleep behavior disorder screening questionnaire; ESS, Epworth Sleepiness Scale; SCOPA-AUT, Scale for Outcomes in Parkinson's disease for Autonomic symptoms; N/A, not applicable; M.I., medication-induced

**Supplementary Table 5. Proportion of cognitive decline** in different monogenic PD forms (according to MDSGene; www.mds.gene.org)

| Gene | Proportion of cognitive decline [%] | Total number |
|---|---|---|
| LRRK2 | 26.7% | n=470 |
| VPS35 | 44.4% | n=9 |
| SNCA | 70.2% | n=94 |
| RAB32 | 36.6%* (35.3% only literature) | n=45* (n=34 only literature) |
| GBA1 | 62% | n=276 |
| PRKN | 19.5% | n=133 |
| PINK1 | 30% | n=110 |
| PARK7 | 35.7% | n=14 |

*including the investigated cohort

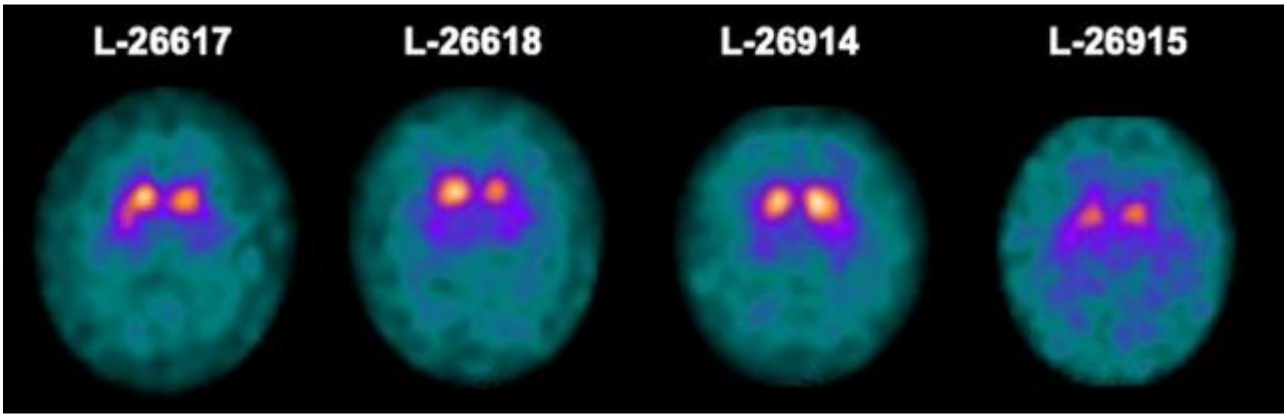


**Supplementary Figure 1.** DaTSCAN Images of patients with PARK-*RAB32*
